# S Gene Target Failure (SGTF) in Commercial Multiplex RT-PCR assay as indicator to detect SARS-CoV-2 VOC B.1.1.7 lineage in Tamil Nadu, India

**DOI:** 10.1101/2021.12.14.21251883

**Authors:** NM Vidya, A Kumaresan, V Kalaivani, A Rajesh Kumar, S Gurunathan, R Avudai Selvi, R Uma, P Varsha, S Raju, P Sampath, T S Selvavinayagam

## Abstract

Emergence of Severe Acute Respiratory Syndrome Corona Virus-2 (SARS-CoV-2) Variants of Concern (VOC) possessing improved virulence, transmissibility and/or immune-escape capabilities has raised significant public health concerns. In order to identify VOCs, WHO recommends Whole-Genome Sequencing approach, which is costly and involves longer completion time. Hence, potential role of commercial multiplex RT-PCR kit to screen variants rapidly is being attempted in this study. A total of 1200 suspected COVID samples from different districts of Tamil Nadu State (India) were screened with Thermo TaqPath RT-PCR kit and Altona’s Realstar RT-PCR Assay kit. Among 1200 screened, S-gene target failure (SGTF) phenomenon were identified in 112 samples while testing with TaqPath RT-PCR Kit. 100% concordant results were observed between SGTF phenomenon and whole-genome sequencing (WGS) results in detecting SARS-CoV-2 VOC B.1.1.7. TaqPath RT-PCR assay testing can be utilized by laboratories to screen rapidly the VOC B.1.1.7 variants, thus enabling early detection of B.1.1.7 variant infection and transmission in population. This in turn will pave way to implement suitable preventive measures by appropriate authorities to control the transmission of the viral variant.

## INTRODUCTION

The current coronavirus disease 2019 (COVID-19) pandemic caused by novel severe acute respiratory syndrome coronavirus 2 (SARS-CoV-2) has affected more than 200 countries globally (Huang et al., 2020; Li et al., 2020; Wu et al., 2020). As on 26^th^ July 2021, 19.5 crore confirmed cases of COVID-19 have been reported worldwide and 41.8 lakh deaths were recorded. At the same time, India recorded 3.14 crore confirmed cases and 4.2 lakh deaths due to COVID-19 (*Https://Www.Worldometers.Info/Coronavirus/*). In India, a medical student travelling from China (Wuhan) on 30th January 2020 was the first reported case of COVID-19 (Vaman et al., 2020).

Over the couple of years, SARS-CoV-2 have evolved to form new variants possessing improved transmissibility, virulence and/or immune-escape capabilities (CMMID COVID-19 Working Group et al., 2021; Davies et al., 2021; Rambaut et al., 2020). Rapid spread of these new SARS-CoV-2 variants poses notable public health challenges (Sánchez-Calvo et al., 2021). As several molecular diagnostics only target small regions in the viral genome, genetic changes in SARS-CoV-2 will significantly affect their diagnosis too (Wollschläger et al., 2021). Among the several SARS-CoV-2 major variants of concern (VOCs) reported till date, B.1.1.7 (UK) is a dominant circulating variant that emerged in England during September 2020 (*Public Health England*, 2021). The new variant has been detected in over 93 countries, including India (O’Toole Á et al., 2021). This VOC has been reported to possess increased mortality and transmissibility (CMMID COVID-19 Working Group et al., 2021; Davies et al., 2021; Leung et al., 2021; Rambaut et al., 2020). Hence, early identification of this variant in patients may help reduce further spread of infection.

Identifying potential B.1.1.7 cases using S-gene target failure (SGTF) results has been recommended by CDC (Galloway et al., 2021). European Centre for Disease Prevention and Control (ECDC) has also suggested laboratories to implement screening RT-PCR tests to detect SGTF among SARS-CoV-2 samples (*ECDC 2021*). However, both CDC and ECDC have emphasized the need to confirm specific variants by sequencing only (*ECDC 2021*; Galloway et al., 2021).

Multiplex PCR is the gold standard for SARS-CoV-2 diagnosis (Kevadiya et al., 2021). Dual or triple targets detection is recommended (Corman et al., 2020; Kevadiya et al., 2021). However, detecting viral variants is not intended with regular PCR kits. Performance of any SARS-CoV-2 assay will be significantly affected by SARS-CoV-2 variants present in samples. Several researchers have recorded gene dropouts using SARS-CoV-2 multiplex PCR approaches targeting S or N or E genes (Artesi et al., 2020; Hasan et al., 2021; Kidd et al., 2021; Umair et al., 2021; Washington et al., 2020; Ziegler et al., 2020). Variant-specific PCR or Whole Genome Sequencing (WGS) techniques are either costly and/or involve longer completion time (Wang et al., 2021; Wollschläger et al., 2021). Availability of a rapid screening approach will enable usage of WGS on samples of interest as well as maximize the use of WGS and expand scope of laboratories to actively participate in VOC identification process.

Thus, the current study focusses on SARS-CoV-2 S gene target failure (SGTF) detection as an indicator for the presence of VOC B.1.1.7 lineage using commercial multiplex PCR assay, further confirmed by whole-genome sequencing.

## METHODOLOGY

### Study Samples, Period and Location

The study was conducted from December 2020 to May 2021 (6 months) involving individuals presenting with ILI symptoms throughout the Tamil Nadu State. Samples from the individuals were classified into one of the following categories *viz*. International Travelers and their close contacts, Community Clusters, Family Clusters, Vaccine Breakthrough Cases, Deceased with no comorbidities, Children and Young Adults with Severe Lung Involvement.

### Sample Collection & Transport

A total of 1200 samples (both oropharyngeal and nasal swabs) were collected in Viral Transport Media from suspected ILI symptoms presenting cases as per the protocol of WHO for sample collection and transport (*WHO 2021*). Care was taken to transport the samples in cold chain to State Public Health Laboratory (SPHL), Directorate of Public Health and Preventive Medicine, Chennai, Tamil Nadu, India as mandated by WHO (*WHO 2021*).

### RNA Extraction

The sample processing was carried out in a Type II Biological Safety Cabinet with HEPA filters, at SPHL, Chennai. RNA was extracted using KingFisher Flex Automated Nucleic Acid Extractor (MVP-II Protocol Kit) as per manufacturer instructions.

### Multiplex Real Time RT-PCR

All the samples were subjected to RT-PCR analysis using TaqPath COVID-19 Combo Kit (ThermoFisher Scientific, USA) that detects 3 genes viz. S, N and ORF-1ab (*ThermoFisher Scientific TaqPath Kit*, 2021). Samples that failed to amplify S-gene in TaqPath COVID-19 Combo kit were subjected to RealStar SARS-CoV-2 RT-PCR Kit (Altona Diagnostics, Germany) testing that identifies E and S genes (*RealStar SARS-CoV-2 RT-PCR Kit*, 2020). The results were analyzed using Quant Studio Design and Analysis Software (v1.5.0). Samples with Cycle Threshold (Ct) value of <25 in RT-PCR were alone selected and sent for sequencing studies.

### Sequencing Studies and Sample Storage

RNA extracts of the samples were sent either to National Institute of Virology (NIV), Pune or to InSTEM, Bengaluru for sequencing studies for confirmation of the variant form of SARS-CoV-2. VTM and excess RNA extracts of the samples were stored at −80°C after RT-PCR analysis. Figure 1 outlines the overview of the study.

**Figure 1.**
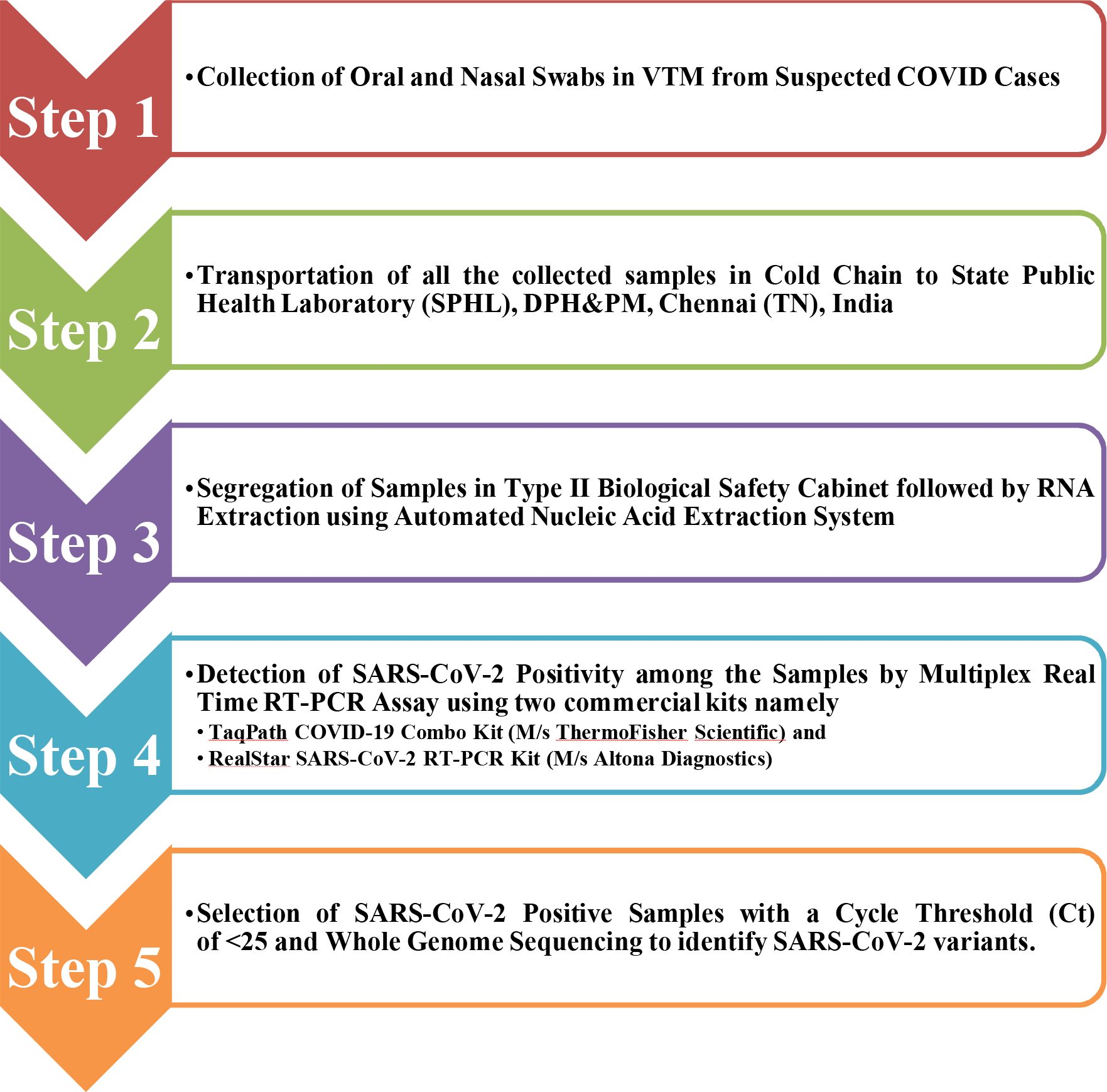
Overview of the Study.

### Ethical Statement

Ethical Approval was obtained for this study from Institutional Ethics Committee (IEC), Madras Medical College, Chennai-3 (ECR/270/Inst./TN/2013/RR-16/No.32092021).

## RESULTS

A total of 1200 respiratory specimens with suspected cases of SARS-CoV-2 were tested by TaqPath COVID-19 Combo Kit. A total of 112 samples among the 1200 samples tested failed to detect S gene in TaqPath COVID-19 Kit during the RT-PCR analysis (Figure 2). However, all these 112 samples reported presence of S gene when tested using RealStar SARS-CoV-2 RT-PCR Kit. All 112 samples reported Ct values of < 25 and hence, were selected for further sequencing studies. 10 samples were sent to National Institute of Virology (NIV), Pune and the remaining 102 samples were sent to InSTEM, Bengaluru. Table 1 displays sequencing results of SARS-CoV-2 Variants among Non-detectable S-gene containing COVID-19 Positive Samples. Sequencing results of 25 samples could not be determined due to insufficient RNA content and too many Ns. In the remaining 87 samples, 75 (86.2%) were identified as Alpha variants (B.1.1.7) followed by 8 samples (9.1%) recognized as Delta variants (B.1.617.2). Only 1.1% samples tested were identified as Eta variant (B.1.525). However, 3.4% of samples were reported to have no variation in their genome content (Ancestral type).

**Table 1:** Distribution of SARS-CoV-2 Variants among Non-detectable S-gene containing COVID-19 Positive Samples (n = 87)

| S. No. | Sequencing Results | No. of Samples | % |
| --- | --- | --- | --- |
| 1 | Alpha Variant (B.1.1.7) | 75 | 86.2 |
| 2 | Delta Variant (B.1.617.2) | 8 | 9.1 |
| 3 | Eta Variant (B.1.525) | 1 | 1.1 |
| 4 | Ancestral | 3 | 3.4 |

**Figure 2.**
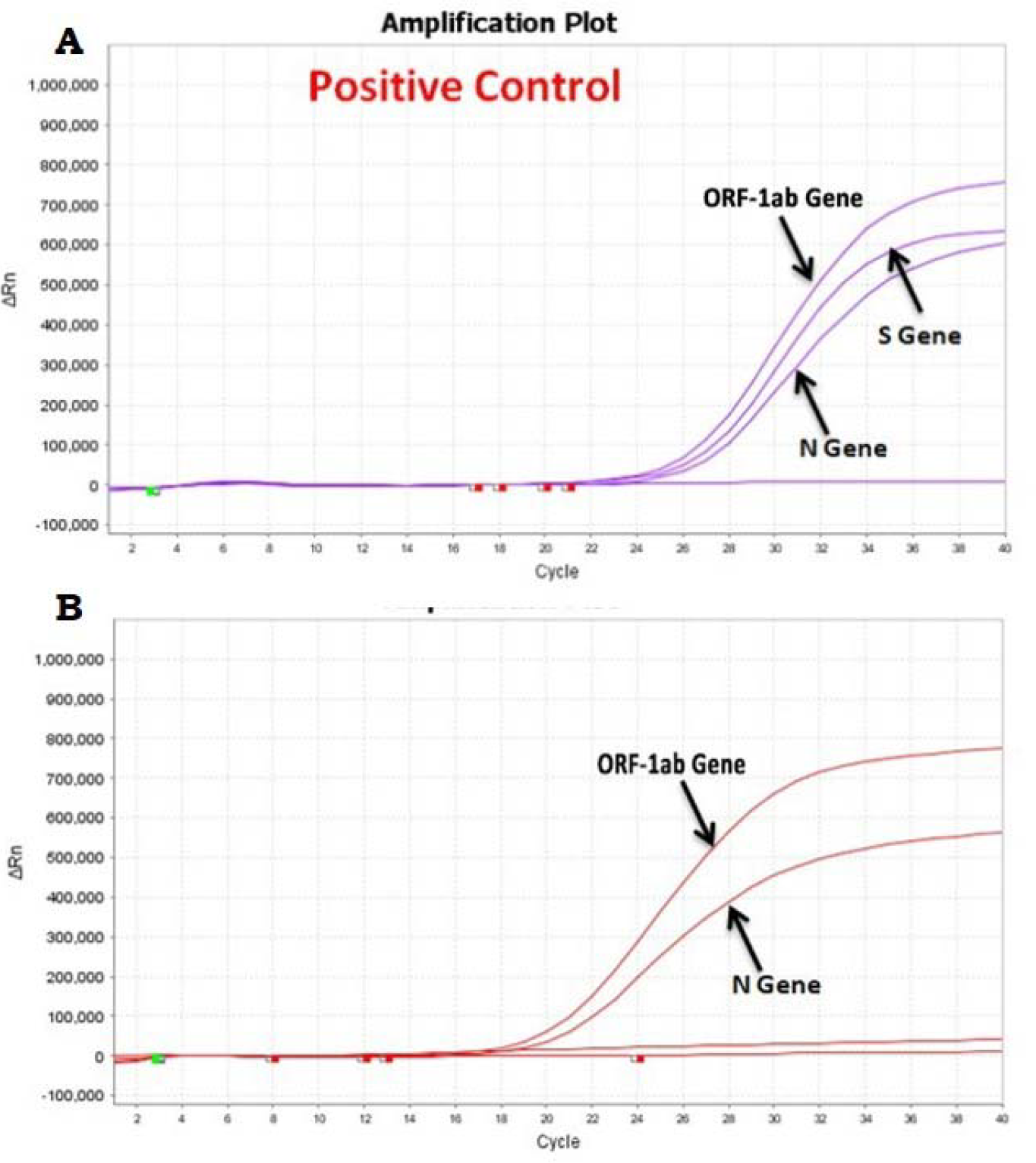
Amplification curves of gene targets with TaqPath RT-PCR Kit (A) Positive Control and (B) Suspected case of VOC B.1.1.7 with S Gene Target Failure (SGTF)

Distribution of SARS-CoV-2 Alpha Variant in the samples analyzed is displayed in Table 2. A majority of the Alpha variants (62.6%) were reported from community clusters (40%) followed by family clusters (23%). In addition, Alpha variants were also reported in 17.3% of the International travelers and their close contact. Alpha variants in the remaining categories were reported to be not more than 8%.

**Table 2:** Distribution of Cases infected with SARS-CoV-2 Alpha Variant (n = 75)

| S. No. | Detail/Description | No. of Samples | % |
| --- | --- | --- | --- |
| 1 | **Category-Wise** |  |  |
|  | Community Clusters | 30 | 40 |
|  | Family Clusters | 17 | 22.6 |
|  | International Travelers and their close contacts | 13 | 17.3 |
|  | Vaccine Breakthrough Cases | 6 | 8 |
|  | Children | 4 | 5.3 |
|  | Deceased | 3 | 4 |
|  | Young Adults with Severe Lung Involvement | 2 | 2.6 |
| 2 | **Age-Wise** |  |  |
|  | <10 years | 5 | 6.6 |
|  | 10 – 18 years | 13 | 17.3 |
|  | 19 - 45 years | 38 | 50.6 |
|  | 46 – 60 years | 13 | 17.3 |
|  | > 60 years | 6 | 8 |
| 3 | **Gender-Wise** |  |  |
|  | Male | 34 | 45.3 |
|  | Female | 41 | 54.6 |
|  | Others | 0 | 0 |

In age-wise distribution, 50.6% of samples among the alpha variants belonged to 19 to 45 age group. 17.3% samples of the alpha variants were reported in two age groups viz. 10-18 and 46-60. Individuals aged above 60 represented 8% of the alpha variants identified, whereas less than 10 years age group formed only 6%. Among the gender-wise distribution, 54.6% and 45.3% of women and men were found to be infected with SARS-CoV-2 alpha variant, respectively.

Table 3 lists month-wise SARS-CoV-2 alpha and delta variants reported during the study period. In April 2021, a maximum of 32 alpha variants (out of 75) and 8 delta variants were recorded (Figure 3).

**Table 3:** Month-Wise Distribution of Cases infected with Alpha and Delta SARS-CoV-2 Variants (n = 83)

| Month/Year | No. of Alpha Variants (n = 75) | No. of Delta Variants (n = 8) |
| --- | --- | --- |
| December 2020 | 03 | 0 |
| January 2021 | 05 | 0 |
| February 2021 | 04 | 0 |
| March 2021 | 17 | 0 |
| April 2021 | 32 | 8 |
| May 2021 | 14 | 0 |

**Figure 3.**
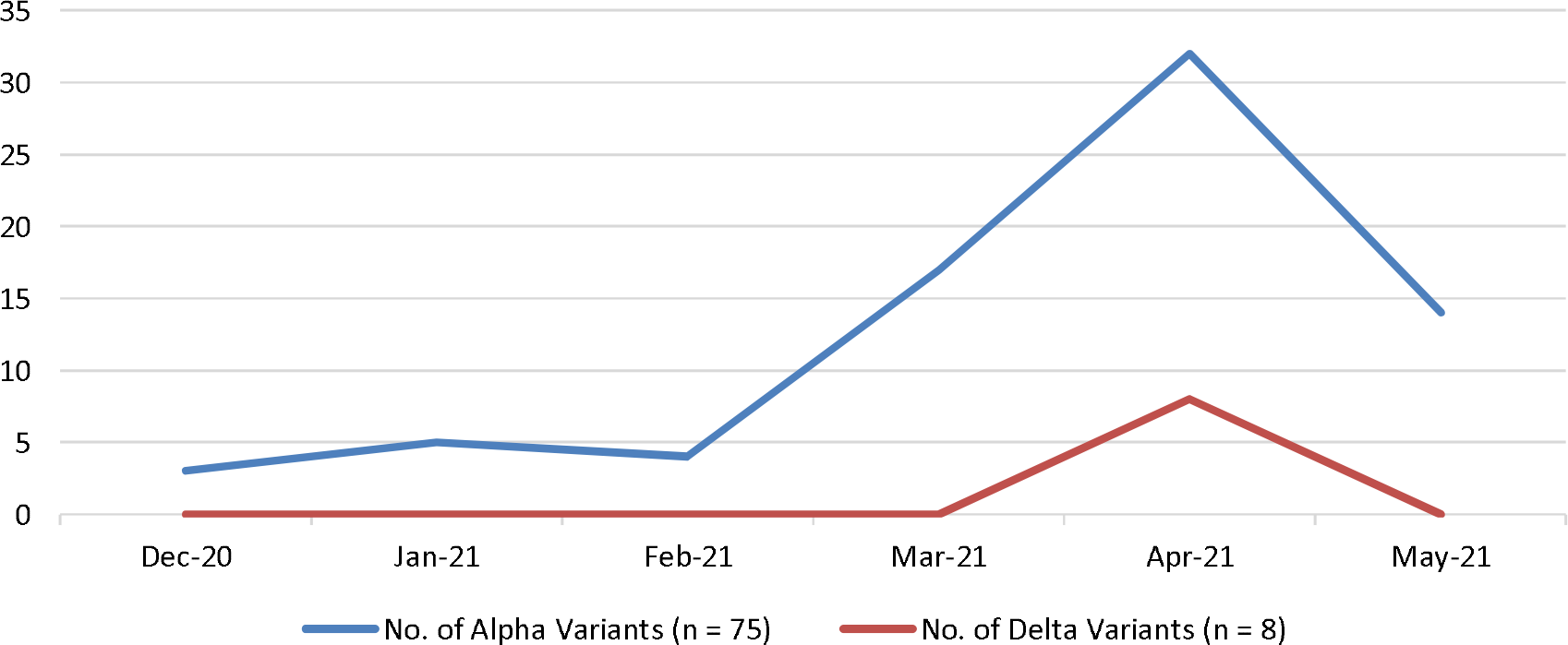
Month-Wise Distribution of SARS-CoV-2 Alpha and Delta Variants in Tamil Nadu.

## DISCUSSION

In the present study, a Multiplex RT-PCR based strategy is demonstrated to reliably screen for VOCs (such as B.1.1.7, B.1.617.2, B.1525) in a laboratory setting. Our results reveal that all the WGS confirmed variants tend to have SGTF by the TaqPath COVID-19 Combo commercial multiplex real time RT-PCR assay (ThermoFisher Scientific, USA). Thus, indicate the potential utility of this screening approach for rapid detection of the VOCs.

The ECDC and CDC recommends timely detection of new SARS-CoV-2 variants as the primary strategy to control the current global pandemic (*ECDC 2021*; Galloway et al., 2021). However, several laboratories consider SARS-CoV-2 variants detection as an additional task, which can be a time bound challenge. Eventhough, WGS is the reference standard to detect, trace and monitor virus variants (surveillance), it is either not accessible to all or easy to perform. TaqPath multiplex assay employing laboratories simultaneously detect SARS-CoV-2 along with the possible detection of VOC. However, laboratories lacking this assay require the re-analysis of the positive samples using another kit/platform (*ECDC 2021*; Galloway et al., 2021; Wollschläger et al., 2021).

Our results report 86.2% samples as Alpha variant (B.1.1.7). Thus, TaqPath assay can be used in places where there is a high prevalence of the VOC B.1.1.7, with little or no co-circulation of other variants that cause other gene dropouts. In this setting, it could be used as a proxy measure of the incidence of this variant. Sánchez-Calvo et al. (2021) have reported similar results based on n-gene drop out or delay.

Among the 1200 samples studied, 112 samples failed to detect S-gene in the TaqPath RT-PCR analysis. However, S-gene was detected when these samples were tested using another commercial multiplex real-time RT-PCR Kit of Altona Diagnostics, Germany (*RealStar SARS-CoV-2 RT-PCR Kit*, 2020). According to CDC and WHO recommendations, S-gene drop could act as an indicator for investigation of new SARS-CoV-2 variant and for further sequencing(Galloway et al., 2021; *WHO 2021*, 2021).

Our study primarily focused on SGTF in SARS-CoV-2 using TaqPath Assay Kit. Kidd et al. (2021), Umair et al. (2021) and Washington et al., (2020) have recorded similar S-gene dropout pattern in SARS-CoV-2. Failure of the S gene target amplification using TaqPath COVID-19 RT-PCR Kit is attributed to H69-V70 deletion typically present in SARS-CoV-2 variant VOC B.1.1.7. Interestingly, other investigators also reported gene target failure phenomenon associated with N and E genes of SARS-CoV-2 (Artesi et al., 2020; Hasan et al., 2021; Sánchez-Calvo et al., 2021; Ziegler et al., 2020). Primary reasons for failure to detect these genes in real-time RT-PCR systems are mutations, deletions or insertions in the gene targets.

From our study, it is clearly evident that Community clusters and Family Clusters in Tamil Nadu were predominantly affected with VOC B.1.1.7 (Table 2). Thus, substantiating increased transmissibility of this variant among the population (Leung et al., 2021). Alpha variant has accounted for higher infections among the 19-45 age group individuals and only 5 cases below age 10 have been reported. Similar results have been reported elsewhere (Davies et al., 2020). The investigators suggest school closure as prime reason for less SARS-CoV-2infection among the children.

Our results reveal peak number of alpha variants reported during April 2021 and also emergence of delta variant infected cases started to be reported at around the same time.

Our study predominantly reports identification of SGTF as B.1.1.7 variant. However, less sample size availability to detect the other two variants during our study is a limitation. Further studies with more samples from diverse population would aid in understanding the newly evolved SARS-CoV-2 infections in our region and its possible selection using a commercially available RT-PCR Assay.

## CONCLUSION

The comparison of Multiplex RT-PCR results by using two different kits targeting different genes allow us to differentiate partial identification of the new variant of SARS-CoV-2 suspicions that is further confirmed by whole genome sequencing results obtained. Multiplex RT-PCR could be used as a potential molecular tool for the rapid identification of SARS-CoV-2 variant so as to ensure efficient screening for timely and accurate diagnosis of SARS-CoV-2 variants. The study findings may aid in risk assessment and will be immediately reviewed to re-assess the overall level of risk to human health and potential implications of SARS-CoV-2 diagnosis, treatment and vaccine development. This will help us to take immediate preventive measures to control the transmission of new variant of SARS-CoV-2 infections in Tamil Nadu state.

## Data Availability

All the data in the present work are provided in the manuscript

## ACKNOWLEDGEMENTS

The authors thank Government of Tamil Nadu, India for supporting State Public Health Laboratory, Directorate of Public Health & Preventive Medicine, Chennai, Tamil Nadu, India. The authors also thank National Institute of Virology (NIV), Pune and InStem, Bengaluru for their support as nodal agency for sequencing of COVID samples.

## AUTHORS CONTRIBUTIONS

Conceived and Designed the Experiments: TSS, SP, SR and NMV. Performed the Experiments: NMV, KA, KV, RKA. Analysed the Data: SR, NMV, UR, VP, SG and RAS. Wrote the Manuscript: SR, NMV, SG and RAS. All authors contributed to data interpretation, critically reviewed the manuscript and approved the final manuscript for submission.

